# Pathways, Perceptions, and the Luck of the Draw: A Qualitative Study of Adolescent Idiopathic Scoliosis Imaging and Referral Services in England

**DOI:** 10.64898/2026.07.16.26358249

**Authors:** Lyn Robinson-Smith, Maya Jafari, Lucksy Kottam, Natalie Clark, Amar Rangan, Joy Adamson, SPINE team

## Abstract

**Introduction:** Adolescent idiopathic scoliosis (AIS) requires frequent x-rays for management, exposing young patients to cumulative radiation risks. While radiation-sparing imaging modalities exist, access across the National Health Service (NHS) remains uneven and information given to patients is variable. This qualitative study investigated the systemic, geographic, and interpersonal dynamics of AIS imaging in England.

**Design:** This qualitative study employed in-depth semi-structured interviews with healthcare professionals (HCPs) from NHS paediatric spinal centres, patients aged 13–25 years old with AIS and parents/carers of young people with AIS.

**Setting:** England.

**Participants:** A total of 22 HCPs from 13/24 NHS paediatric spinal centres in England, 19 10-25 years with AIS and 11 parents/carers.

**Results:** Conventional x-ray remains the main imaging modality. Significant geographic inequality exists. The most commonly available radiation-sparing imaging modality available is the EOS system, which uses slot-scanning technology, is available at 7 centres in England, primarily in London imaging networks. Acquisition of EOS systems is currently driven by local charitable funding rather than a centralised strategy, with high capital and installation costs cited as primary barriers. Inconsistent knowledge of imaging within primary care and a lack of specialist expertise in local secondary care services led to diagnostic redundancy, gatekeeping, and low value inconsistent imaging. These systemic delays frequently closed the window for conservative treatments like bracing. A professional balancing act exists between the duty to inform and the desire to minimise patient anxiety. HCPs often use selective communication regarding radiation risks. Conversely, families demonstrate high relational trust with HCPs and low baseline knowledge of cumulative exposure, often viewing frequent imaging as a reassuring marker of clinical progress. In centres with EOS systems, clinicians felt empowered to lead proactive, transparent risk discussions. In standard X-ray settings, dialogue remains reactive and infrequent, leading to a reliance on implied rather than truly informed consent.

**Conclusions:** AIS imaging in England is variable. Geographic location dictates access to low-dose radiation technology and the quality of informed consent. Systemic inefficiencies and fragmented referral pathways contribute to diagnostic redundancy and delayed specialist care. National standardisation of clinical pathways, information provision and a centralised strategy for low-dose technology procurement are essential to eliminate structural inequalities and ensure equitable, transparent care for all patients.

**STRENGHTS AND LIMITATIONS OF THIS STUDY:**

- **Multidisciplinary and multi-perspective Insight:** The study captured a comprehensive overview of the AIS landscape by collecting and analysing data from 22 healthcare professionals representative of different roles within the NHS delivering specialised AIS care alongside the lived experiences of 30 patients and parents/carers.
- **Rigorous co-design and patient and public involvement (PPI):** The methodology was robustly informed by a Patient Advisory Group (PAG) and a PPI co-applicant, ensuring that interview topic guides and recruitment materials were child-friendly, age-appropriate, and focused on outcomes meaningful to the AIS community.
- **Adherence to reporting standards:** Transparency and methodological rigour were maintained by following the Standards for Reporting Qualitative Research (SRQR) guidelines and employing Braun and Clarke’s [8] established six-phase thematic framework for systematic data analysis
- **Digital recruitment bias:** Recruitment of patients and carers was primarily conducted via social media platforms, which may have excluded individuals with limited digital literacy or those not engaged with the Scoliosis Support and Research (SSR) network, potentially limiting the diversity of the sample.
- **Lack of ethnic diversity:** The sample lacked sufficient representation from minority ethnic groups in the patient group; as AIS prevalence and treatment outcomes can vary across different backgrounds, the findings may not fully reflect the specific cultural or systemic barriers faced by these populations.

## INTRODUCTION

Scoliosis is a 3-dimensional deformity of the spine. It affects up to 5.2% of adolescents between the ages of 10-18 years. No cause is found in 80% of cases, and this is called ‘Adolescent Idiopathic Scoliosis’ (AIS). Imaging plays a central role in the diagnosis and monitoring of AIS. The most common modality used for AIS imaging is conventional radiography (x-rays). However, radiography uses ionising radiation, which carries an inherent risk of causing DNA damage [1]. This risk is of particular relevance in younger patients, who have a higher proportion of actively dividing cells and a longer lifetime during which radiation-induced effects may manifest [2]. Although the absolute risk from individual x-ray exposures is low, there are existing concerns relating to cumulative radiation exposure. This is especially pertinent for AIS patients, where repeated imaging is a routine component of care. Many AIS patients undergo approximately 10–25 radiographs during adolescence, and some receiving 40–50 or more in cases requiring prolonged or complex monitoring [3]. Research shows that healthcare professionals tend to have a limited understanding of radiation risk and are therefore poorly equipped to correctly inform patients about imaging-related risks [4] and we know little of how much information is shared with AIS patients regarding radiation risk. Whilst a range of radiation-sparing scoliosis imaging modalities (such low-dose slot-scanning technology like EOS) exist [5–7], there is a lack of knowledge as to their provision within National Health Service (NHS) paediatric spinal centres in England.

The Scoliosis Imaging Needs Patients Evaluation (SPINE) study included a qualitative component, the primary objective of which was to map current AIS imaging service provision in England and identify areas for improvement. Data were collected through semi-structured interviews with key stakeholders. Through analysis of these data, the study aimed to: (i) explore stakeholder perspectives on current AIS imaging services, including modes of delivery and the information provided to patients and their families; and (ii) examine how imaging-related risk is communicated in clinical practice, the implications of variation in this communication, and areas requiring improvement and greater consistency.

## METHODS

### Study Design

This qualitative study comprised in-depth semi-structured interviews to gain the insights of HCPs, patients with AIS, and parents/carers. A topic guide was developed which included open-ended questions. The SRQR guidelines were used where applicable. The study was co-designed and conducted by a research team that included academic, clinical and PPI representatives.

### Sampling and recruitment

#### Healthcare professionals

Initially, purposive recruitment was initiated through established contacts at England based paediatric spinal centres and the NHS England Neurosurgery and Spinal Surgery Clinical Reference Group (CRG) and the British Scoliosis Society (BSS). The study was advertised through member forums of the British Society of Skeletal Radiologists (BSSR), and the British Society of Paediatric Radiologists (BSPR). To further broaden the sample, paediatric spinal surgeons in underrepresented centres were invited via email. Invitations were distributed through the allied health professional group of the BSS. Snowball sampling was implemented to identify further multidisciplinary stakeholders with specialised expertise in the area.

#### Patients, Parents and Carers

Patients, parents and carers were recruited via Scoliosis Support and Research (SSR), which advertised the study via Facebook (over 18,000 followers) and Instagram (over 11,000 followers), study-specific social media pages and collaborative networks. Interested individuals were requested to read an online participant information sheet, and if they were happy to participate, completed an online consent form. Recruitment thereafter followed an iterative purposive sampling approach to include participants with diverse characteristics. While the recruitment was primarily restricted to social media users, the membership breadth was sufficient to achieve the required clinical variation to meet the study’s aims.

### Data collection

Semi-structured interviews were conducted via videoconferencing or telephone, based on participant preference and logistical considerations. Tailored topic guides were developed for each group of stakeholders which were informed by consultations with the research team and PPI representatives. Interviews were designed to last approximately 45 minutes for HCPs and 30 minutes for patients and parents/carers. As a token of appreciation, patients and parents/carers received a £15 gift voucher. With participant informed consent, all interviews were audio-recorded and transcribed verbatim for analysis.

### Analysis

Data organisation and retrieval were facilitated using NVivo software. Data were analysed thematically according to the six-phase framework established by Braun and Clarke [8]. Transcripts from interviews helped to create a descriptive overview of service delivery and patient pathways across England, highlighting variations between NHS sites. Initial qualitative analysis was conducted independently for healthcare professionals and then patients and parents/carers, followed by intra- and inter-group comparisons. While some themes were group-specific, overarching themes were developed. To ensure rigour, code and theme development were reviewed through regular team debriefs throughout the analytical process.

### Public and patient involvement

The study’s PAG included 6–8 members with lived experience of Adolescent Idiopathic Scoliosis (AIS) and their families, alongside a PPI co-applicant within the Trial Management Group. The PAG actively informed the study’s lifecycle:

#### Design & Materials

They prioritised research topics and co-developed information sheets and topic guides. A Year 6 primary school class also helped create child-friendly materials to ensure accessibility.

#### Analysis

Members provided patient perspectives to help interpret emerging findings.

#### Dissemination

The PAG and PPI co-applicant co-produced lay summaries, academic outputs, and social media content, while advising on effective ways to engage professional societies and patient support groups.

## RESULTS

### Participants

#### Healthcare professionals

Twenty-two HCPs from 13 of the 24 NHS paediatric spinal centres in England participated in semi-structured interviews (19 videoconferencing; 3 telephone). To ensure maximum variation, a diverse range of roles were recruited, including paediatric spinal surgeons, nurses, radiologists, radiographers, radiology service managers, orthotists, and physiotherapists (8 male; 14 female). Of the 13 centres represented, four utilised EOS systems. To maintain participant anonymity, specific recruitment sites are not disclosed.

#### Patients

Nineteen young people, aged 13-25 years of age (mean age 18) participated in interviews (11 videoconferencing; 8 telephone). No participants from the youngest targeted demographic (10-12 years old) were recruited. This discrepancy likely stems from the recruitment strategy’s reliance on social media, where younger children often face restricted independent access. The sample was predominately female (17 female 2 male), receiving care across 11 of the 24 specialist spinal centres in England. The clinical characteristics of the cohort were that 13 had undergone surgery (three had braced prior to surgery), three were bracing at the time of interview, and three “watch and wait” observation. We have withheld describing clinical characteristics by gender to preserve anonymity. This gender distribution aligns with the known epidemiology of AIS; females are 5–10 times more likely than males to develop scoliosis and exhibit a significantly higher risk of progression to severe curvature [9]. Consequently, these clinical trends are appropriately reflected within the current study sample. The sample lacked ethnic diversity, with all patient participants identifying as White.

#### Parents/carers

Eleven parents/carers participated in interviews (7 videoconferencing; 4 telephone), representing 7 spinal centres and 1 local hospital. The sample was predominantly female (10 female; 1 male). Notably, interview participation was not contingent on child enrolment; 5 parents/carers participated independently of a child’s direct involvement in the study.

### Thematic analysis

#### Theme 1 – Service infrastructure and the availability of imaging modalities

Conventional radiography (x-ray) remains the standard imaging modality for diagnosis and monitoring of AIS, with a typical six-monthly review, across sites. However, frequency of clinical review and repeat imaging varied significantly depending on the age, gender and severity at which the curve was identified. Data revealed significant geographic inequalities regarding access to radiation-sparing AIS imaging modalities. Surface topography was available and utilised in only one centre within our sample, and computed tomography was predominantly used in pre-operative planning and to look for post-surgical complications. At the time of study, EOS imaging was available at only seven NHS paediatric spinal centres in England, with a concentration in London imaging networks. Furthermore, accessibility was constrained by integrated care models; healthcare professionals noted that patients could not travel to a different centre solely for low-dose imaging, as the entire multidisciplinary care pathway for AIS was required to remain at a single site. Participants generally lacked awareness of radiation-sparing systems, most did not acknowledge radiation as a variable factor in their care. The minority who were informed acknowledged the significant geographic and pathway-related constraints:

> *“EOS would be better… but the access is very, very limited. If* [patient name] *would have turned up at* [a centre with EOS] *and said, “the doctor sent me for a spinal x-ray, but I know you’ve got EOS and that’s better for spines”. They would have been like, “well, you’ve not been referred for EOS here, you’ve been referred for x-rays, so you we can’t do it”*. (Parent/carer)

The initial acquisition of these systems was described by HCPs as taking multiple years, most often dependent on successful local charitable funding bids and reallocation of department funds, rather than centralised clinical strategy. Several HCPs reported that they had submitted unsuccessful business cases for EOS systems. Hospital administrations frequently declined business cases for low-dose imaging systems, citing a combination of prohibitive initial capital expenditure, installation costs, and/or the high cost of ongoing service contracts. These financial barriers were often compounded by structural constraints, such as insufficient floor space and/or power, and strategic challenges, including competing institutional priorities. Furthermore, in some trusts, the specific paediatric population was deemed too small to provide a clinical return on investment that justified the substantial costs of the system. One centre explained:

> *“Great efforts have been put in to try and get one* [an EOS], *and we’ve been utterly unsuccessful. One of my colleagues has been instrumental in trying to work very hard for several years to get the machine, on the basis of reduced radiation exposure for children, but it’s not been possible to get one due to several issues really. There’s the overall cost. However, we that got round it* [by securing charity funding]. *The radiology department weren’t too happy having a machine that could only really mainly be used to spinal X-rays rather than everything else, taking some valuable space. But then we did manage to then find a place in the hospital that could go near our clinic that was going to work. The actual stumbling block was the £300,000 labour costs to install it. So, whilst we could find money from somewhere for the actual machine, installing it, strengthening the floor…whatever they have to do, killed it. Huge efforts had been made; architects had made plans*. [Colleague] *was heartbroken about it…*.*the amount of effort he’s put in to try to get this machine. And yet we haven’t got one yet. We then continue to expose you into more radiation than we need to”* (HCP).

#### Theme 2 – Clinical pathways and impact on radiation exposure

The imaging pathway was frequently complicated by systemic inefficiencies and diagnostic redundancy. The patient journey typically started in primary care, where GPs demonstrated varying and often limited knowledge regarding AIS. In certain regions, GPs were permitted to request X-rays for suspected AIS, a practice that frequently delayed the formal referral process and increased the prevalence of clinically inadequate imaging. Some GPs ill-advised “watch and wait” strategies. Most patients were referred to either local secondary care services or specialist paediatric spinal centres, depending on their locality. Local secondary care services often possessed limited specialised expertise in AIS management and subsequently, inconsistencies were found in monitoring frequency, protocols and quality of referrals to specialist paediatric spinal centres. These delays heightened the risk of patients presenting to specialist care with severe curvatures, at which point conservative treatment options were no longer viable.

“*I think we’re seen as a surgical service by some consultants* [at secondary care hospitals] *and therefore they refer them when they want surgery. And actually we’re the bracing providers too…. We would have liked to prevent the surgery if we could have done so. Yeah, we definitely want early referrals, but we don’t always get them”* (HCP).

Initial encounters at primary and local secondary care services often set a trajectory that was layered, repetitive and one of gatekeeping rather than a streamlined service. Many families required a second opinion or private consultation to access specialist review. A recurring theme in the data was the necessity of repeat X-rays upon a patient’s transition to specialist care. This occurred because previous radiographs were either technically sub-standard for clinical requirements, clinically outdated due to referral delays, or had not been successfully transferred between NHS trusts prior to the initial appointment. Consequently, the absence of seamless data integration and early specialist oversight resulted in an increased cumulative radiation burden for some young patients.

Wait times within specialist paediatric spinal centres exhibited significant geographic variation. While clinical staff reported that severe cases were escalated up the referral pathway quickly upon identification, patient processing protocols, conservative care and waiting times for operations varied considerably. There was a lack of consistency in service delivery models between different spinal centres. Patients and their parents/carers often characterised this systemic variability of patient pathways as being determined by luck rather than a lack of standardised clinical protocols.

> *“Every moment it was luck… I don’t have a lot of faith in the system. I feel like “lucky us” that we wriggled through it and got where we got. It could have been very different”* (Parent/carer).

#### Theme 2 – Risk communication in AIS imaging

Healthcare professionals demonstrated an awareness of the risks associated with cumulative radiation exposure in AIS monitoring, however, struggled to quantify these risks with precision as imaging frequency was highly individualised. Healthcare professionals found it difficult to be fully transparent because they lacked exact data on long-term outcomes and struggled to translate radiation risks into everyday language for families; many reported having had limited training on the matter. Consequently, a shared professional consensus emerged that the immediate clinical benefits of accurately monitoring curve progression to inform care outweighed risks of radiation exposure. In this context, frequent imaging is framed as an essential component of quality care:

> *“What is the risk of radiation? I don’t know. Because there are many factors involved… How many X-rays? What are the strength of X-rays used? But the more important question is, would it change the management? I don’t think so…* [healthcare professionals] *request the bear minimum X-rays needed. I don’t think it would change the management as such*… *Yes, there is a risk, but there’s a risk of not doing X-rays as well”* (HCP).

This professional perspective necessitated a balancing act. Navigating a tension between the ethical obligation to provide informed consent and a competing duty to minimise unnecessary patient anxiety. This was particularly evident in cases where imaging was deemed a clinical necessity and imaging options could not change regardless of the discussion. Many HCPs acknowledged a strategy of selective communication whereby; detailed risk discussions did not take place unless initiated by the patient/family. Rather than a failure of transparency, HCPs wanted to avoid creating fear around a mandatory clinical pathway. There was a prevailing view that emphasising a statistically small risk might undermine a family’s confidence in a necessary treatment plan without offering a viable alternative.

“*So when I do X-rays, what I say to them is… “look, there’s always a risk and a balance between everything… the risks aren’t hugely high and I can’t give you a number about what your risk would be, but we know that the less X-rays we take the better. We’re going to take as few as we can get away with, but as many as we need. And if there’s a chance to start stretching that time frame out, then we’ll stretch it out”. So that’s kind of the discussion I have with them*”. (HCP)

There was a lack of clarity around who was responsible for discussing cumulative radiation risk with AIS patients and their families. Whilst the administrative task of obtaining formal consent typically fell to radiographers at the point of imaging, it was recognised as a suboptimal setting for addressing the complexities of cumulative radiation exposure. Spinal consultants were viewed as having the necessary clinical context for such discussions however their ability to facilitate these was constrained by systemic pressures including limited consultation windows and extensive waiting lists. These factors marginalised radiation risk dialogue within the standard care pathway.

Notably, the availability of advanced low-dose imaging technology such as EOS significantly altered the communicative dynamic. HCPs in centres with low-dose systems reported feeling empowered to discuss radiation risk proactively. The presence of a ‘safer’ alternative to standard x-ray transformed the conversation from one of risk mitigation to one of technological advantage. The availability of EOS saw HCPs addressing radiation concerns without patient prompts, fostering a more open dialogue that was often absent in sites that only had standard x-ray.

> *“I’ve heard my consultants say this* [EOS] *is 10 times less radiation than your standard X-ray. So before you could have been exposed to the full traditional radiation level. You would have had to have 10 of these. So when they* [patients and families] *hear that, they feel a lot more comfortable and comforted by the idea of regular scans, in order to be able to monitor the risk of curve progression, and we can easily justify why we’re doing them” (*HCP).

Participants recalled receiving minimal to no information concerning radiation risk - “*they never talked about x-rays. It was just a thing that I had to do*” (Patient). Several patients and most parents/carers had a very basic underlying awareness of radiation risk and most viewed imaging metaphorically; “*a very small part of a very big jigsaw “* (parent/carer). Families relegated concerns about radiation and prioritised the urgent clinical management of the condition.

Trust in clinical expertise acted as a primary mediator; families operated under the assumption that if an image was requested, the benefit outweighed any risk. Imaging was a primary marker of clinical progress for families and the omission of imaging during a consultation was a source of significant anxiety. HCPs frequently found themselves in the paradoxical position of having to justify the absence of imaging and complicated their efforts to minimise exposure. Regular radiography was socialised into the care pathway not only as a diagnostic tool but as a form of clinical reassurance. Several HCPs expressed a sense of professional guilt regarding their routine non-disclosure of cumulative radiation risks. They expressed a desire for standardised, national information resources (such as co-designed patient leaflets or infographics) to ensure consistent risk communication and facilitate informed consent across all spinal centres.

“[x-ray is an] *investigation based on what we call implied consent, whereas if they don’t object to it, it’s OK. But how can they object to it if they don’t know what the problem is?*… *Most of the time, people just do what you tell them, and I think we’re probably not giving them very much informed consent*.” (HCP).

## DISCUSSION

This study identifies a fragmented imaging landscape for AIS patients in England, where a patient’s journey is often defined by geographic variation rather than standardised clinical pathway. The principal findings reveal that access to low-dose EOS technology is concentrated in imaging networks within London and often dependent on local charitable funding. Furthermore, a professional balancing act exists between the duty to inform regarding radiation risk and the desire to minimise patient anxiety. Clinicians often employ selective communication to maintain patient reassurance, while families often view regular imaging as a primary marker of high-quality care and clinical progress. The presence of low-dose radiation imaging such as EOS facilitated proactive risk communication around cumulative radiation exposure between professionals and patients.

These findings extend the existing research on NHS postcode lotteries [e.g. 10-12] by illustrating how regional inequalities manifest through both physical access to technology and the quality of clinical communication. Ultimately, a patient’s geographic location serves as a primary driver of inequality, influencing not only the cumulative radiation burden they receive but also the streamlining of their referral pathway and their ability to engage in informed decision-making.

The implications for clinicians, service managers and policymakers are significant. Inefficiencies in cross-Trust image sharing frequently meant duplicate imaging, increasing the cumulative radiation exposure for patients, while introducing avoidable delays within the specialist referral pathway. These findings reinforce wider NHS ambitions around digital transformation and ‘making data work harder’ [13], particularly around imaging networks that should allow timely transfer and review of imaging [14,15]. There is currently variation in access to radiation-sparing AIS imaging modalities and inconsistencies in referral pathways. GPs’ restricted access to AIS imaging in some geographical areas and limited AIS expertise in primary and local secondary care services appear to contribute to gatekeeping behaviours that delay specialist referrals. These delays often mean young people with AIS present later to specialist centres and are less likely to be suitable for non-surgical management.

The current system relies heavily on relational trust, which, while protective against immediate anxiety, creates a systemic gap in informed consent and technological equity. At the time of writing, only 7 centres in England had EOS systems, with even fewer centres having access to other radiation-sparing AIS imaging modalities, like surface topography and dual-energy X-ray absorptiometry. The UK National Institute for Health and Care Excellence (NICE) guidance on EOS system was informed by the last systematic review and economic analysis on EOS systems conducted in 2011 [16–17]. Since then advances in slot-scanning technology, including development of the microdose-EOS, with further reduction in radiation exposure have expanded the available evidence base relating to this technology [18–19]. In light of these developments, and as acknowledged by South et al. [20], it is timely to undertake an updated systematic review and economic evaluation of EOS systems and other slot-scanning technologies in AIS imaging. Policymakers should consider a centralised clinical strategy for the procurement of low-dose imaging to eliminate geographic disparities. Furthermore, HCPs require standardised, age-appropriate frameworks to discuss cumulative risk confidently, moving away towards a model of informed confidence.

### Strengths and limitations of this study

This qualitative AIS study maintained methodological rigour by adhering to SRQR guidelines, utilizing Braun and Clarke’s [8] framework, and embedding a PAG to ensure child-friendly materials. A major strength is its dyadic approach, combining clinician perspectives with patient-family units to offer a holistic view of the care pathway. However, while capturing deep regional experiences, these findings may not reflect the operational nuances of every NHS Trust. Furthermore, digital recruitment via social media and the SSR network potentially excluded individuals with limited digital literacy. Lastly, the authors recognise that greater diversity regarding gender and ethnicity would have enhanced the breadth of perspectives, meaning the findings may not fully capture the specific cultural or systemic barriers faced by minority populations.

## CONCLUSIONS

This study exposes a fragmented English AIS imaging landscape, where geographic location structurally determines a patient’s cumulative radiation burden. Procurement of EOS low-dose technology relies on local charitable funding rather than centralised NHS strategy, access remains highly inequitable. Primary and secondary care gatekeeping causes diagnostic redundancy and referral delays, often closing the window for conservative bracing. Furthermore, reliance on high relational trust and selective risk communication masks a systemic gap in truly informed consent regarding x-ray monitoring. Conversely, low-dose technology acts as a powerful clinical mediator, empowering practitioners to engage in proactive, transparent risk communication. To ensure equity, a centralised procurement strategy for radiation-sparing imaging systems must be implemented via NHS England frameworks. National guidelines must also standardise referral pathways and risk communication. Future research should investigate how selective imaging risk communication impacts patient autonomy during adult transition and explore the development of a unified national AIS pathway.

## DATA AVAILABLILTY STATEMENT

Due to the ethical restrictions and the identifiable nature of qualitative interview data involving young patients and healthcare professionals, the full dataset cannot be made publicly available. Requests for access to anonymised data fragments can be directed to the corresponding author at the University of York for researchers who meet the criteria for access to confidential data.

## ETHICS STATEMENTS

### Patient consent for publication

Not applicable.

### Ethics Approval

This study was approved by the Health Sciences Research and Governance Committee at the University of York (HSGRC reference HSRGC/2025/688/H). Informed consent was gained from participants before taking part.

## ACKNOWLEDGEMENTS

We would like to sincerely thank all participants who generously gave their time to take part in this study and the members of the patient advisory group for their contributions. We are particularly grateful to the young people living with adolescent idiopathic scoliosis and their families for sharing their personal experiences so openly. Their insights have been invaluable in helping to better understand AIS care and inform improvements in practice.

On behalf of the SPINE Team

Emily Cooper (Patient and Public Representative), Steven Liggert^1^, Emily South^2^, Ros Wade^2^, Melissa Harden^2^, Matthew Walton^2^, Jiongyu Chen^2^

^1^South Tees Hospitals NHS Foundation Trust, Marton Road, Middlesbrough, UK

^2^Centre for Reviews and Dissemination, University of York, York, UK

